# Improving rational medicine prescribing at a public district hospital in southwestern Uganda, January–November 2021: a quality improvement initiative

**DOI:** 10.64898/2026.09.02.26361584

**Authors:** Richard Migisha, Sharon Namasambi, Benon Kwesiga, Lilian Bulage, Alex Riolexus Ario

**Author notes:** **Corresponding author** Richard Migisha, Uganda Public Health Fellowship Program, Uganda National Institute of Public Health, Ministry of Health, Kampala, Uganda.

## Abstract

Irrational prescribing can compromise patient safety, medicine availability, and antimicrobial stewardship. At a public district hospital in southwestern Uganda, a 2019 internal audit prompted by recurrent stock-outs found approximately three medicines per outpatient prescription, four per inpatient prescription, and antibiotics in about 60% of outpatient prescriptions. We implemented a six-month quality improvement initiative to increase prescribing concordant with the Uganda Clinical Guidelines (UCG) and improve key indicators of rational medicine use among outpatients. From January to November 2021, we conducted an uncontrolled before-and-after quality improvement evaluation. Prescription records were selected by systematic random sampling at baseline and six-month endline. We reviewed 350 baseline and 101 endline outpatient prescriptions. The primary outcome was prescription concordance with UCG. Secondary outcomes included antibiotic prescribing, number of medicines per encounter, generic prescribing, and injection use. Adjusted analyses accounted for age, sex, and diagnostic group. Following root-cause analysis, the hospital quality improvement (QI) team implemented prescriber education, provided clinicians with soft-copy PDF versions of the UCG and essential medicines lists, restricted prescribing to recognized clinicians with signature verification, and instituted periodic prescription audit and feedback. Guideline-concordant prescribing increased from 43.7% to 76.2% (adjusted change, +32.8 percentage points; 95% CI, 21.9–43.8; aPR=1.82; 95% CI, 1.51–2.20). Antibiotic prescribing decreased from 83.4% to 65.3% (adjusted change, −17.9 points; 95% CI, −27.8 to −8.0; aPR=0.80; 95% CI, 0.70–0.91). Mean medicines per encounter decreased from 3.35 to 2.66 (adjusted mean difference, −0.61; 95% CI, −0.79 to −0.43), while prescriptions containing only generic medicines increased from 78.6% to 94.1% (adjusted change, +13.6 points; 95% CI, 5.8–21.4). Guideline-concordant prescribing increased, while antibiotic use and medicine burden declined and generic prescribing improved. Antibiotic use nevertheless remained high, underscoring the need for continued stewardship, audit, and iterative QI.

## Introduction

Rational prescribing is central to patient safety, medicine availability, and efficient use of health-system resources. The World Health Organization (WHO) defines rational medicine use as providing patients with medicines that meet their clinical needs, at appropriate doses and duration, and at the lowest cost to patients and communities [1]. Yet inappropriate medicine use remains widespread, commonly manifesting as polypharmacy, unnecessary antimicrobial prescribing, excessive injection use, failure to follow treatment guidelines, and use of unnecessarily expensive medicines [1]. In resource-constrained settings, these practices may compound medicine shortages, increase avoidable treatment costs and adverse effects, and accelerate antimicrobial resistance. WHO and the International Network for Rational Use of Drugs developed standardized prescribing indicators to enable health facilities to identify and monitor such prescribing gaps [2].

These prescribing challenges are particularly important in settings where medicine supply, diagnostic capacity, and clinical staffing are already constrained. Across 43 studies from 11 countries in the WHO African Region, the median number of medicines prescribed per encounter was 3.1, almost half of encounters included an antibiotic, and only 68.0% of medicines were prescribed generically [3]. Uganda has shown comparable gaps. In the national SPARS baseline assessment of 1,384 health facilities, prescribing quality was the lowest-performing medicines-management domain[4]. More recently, only 34.2% of nearly 10,000 antibiotic prescriptions in lower-level facilities in southwestern Uganda were concordant with the Uganda Clinical Guidelines (UCG), and approximately half were written by cadres not authorized to prescribe[5]. High antibiotic use has also been reported elsewhere in the country [5–7]. As the UCG provide standardized, evidence-based guidance for the management of common conditions, improving concordance with these guidelines represents an important target for facility-level quality improvement [8].

At Itojo Hospital, a public district hospital in southwestern Uganda, recurrent medicine stock-outs during 2018–2019 raised concern that potentially avoidable medicine use might be placing additional pressure on limited supplies. A 2019 internal hospital audit found an average of approximately three medicines per outpatient prescription and four per inpatient prescription, while about 60% of outpatient prescriptions included an antibiotic, identifying rational prescribing as an important target for quality improvement. Previous work in Uganda has shown that educational and guideline-based interventions can improve prescribing, although sustained improvement may require approaches that extend beyond education alone [9]. Uganda’s national SPARS experience likewise supports combining standardized tools, supervision, performance measurement, and feedback to strengthen medicines management and prescribing practices[4]. We therefore undertook a facility-led quality improvement initiative to identify modifiable contributors to inappropriate prescribing and implement locally appropriate changes. The specific aim was to increase the proportion of prescriptions concordant with the Uganda Clinical Guidelines by 30% within six months while improving other indicators of rational medicine use. Guideline concordance was the primary outcome, with antibiotic prescribing, number of medicines per encounter, generic prescribing, and injection use assessed as secondary outcomes.

## Methods

### Study design and setting

We conducted a single-facility, uncontrolled before-and-after quality improvement (QI) evaluation at Itojo General Hospital in Ntungamo District, southwestern Uganda, from January to November 2021. Itojo is a government general hospital providing inpatient and outpatient services, with an estimated bed capacity of approximately 120 [10].

The QI initiative focused on prescribing in the general outpatient department (OPD) and was undertaken in response to recurrent medicine stock-outs and concerns raised by a 2019 internal prescribing audit. The 2021 baseline assessment formed the initial measurement phase of the QI process. The specific improvement aim was to increase the proportion of prescriptions concordant with the Uganda Clinical Guidelines (UCG) by 30% within six months while improving other indicators of rational medicine use.

### QI team and development of the change package

An inception meeting was held with the Ntungamo District Health Office and Itojo Hospital leadership to obtain institutional support. The existing hospital QI team was expanded to include medical, clinical, nursing, pharmacy, dispensing, laboratory, and administrative staff, with the clinical head of the OPD serving as team mentor. Team members received training in QI methods, including development of improvement aims and measures, selection and testing of change ideas, and Plan-Do-Study-Act principles.

Following the baseline assessment, the team reviewed identified prescribing gaps and used fishbone analysis, the five-whys technique, and process mapping of the prescribing and dispensing pathway to identify potentially modifiable contributors.

### QI interventions

From May through November 2021, the hospital implemented a multifaceted change package informed by the root-cause assessment. Interventions included prescriber education and continuing medical education on rational medicine use; provision of soft-copy PDF versions of the UCG to OPD clinicians; provision and periodic updating of essential medicines lists; restriction of prescribing to recognized clinicians; provision of authorized prescriber signatures to pharmacy and dispensing staff; and periodic prescription audit and feedback.

The components were implemented as a facility-level package within the same improvement period; therefore, the evaluation assessed overall change during the QI period rather than the contribution of individual intervention components. Quantitative measures of intervention fidelity, including training attendance, measured guideline availability, and the number and timing of audit-feedback cycles, were not sufficiently complete for retrospective analysis.

### Prescription sampling and data collection

The unit of analysis was an outpatient prescription encounter. Routine OPD prescription and dispensing records were reviewed before and after implementation, with eligible records selected using systematic random sampling at both assessments.

For the baseline assessment, records covering January–March 2021 were reviewed to obtain a broad sample of routine pre-intervention prescribing. A total of 350 prescription encounters were systematically sampled. Endline prescribing was assessed over a three-week period in November 2021, approximately six months after initiation of the QI interventions. After application of the data-quality procedures described below, 101 endline prescription encounters were included in the primary analysis.

For each sampled encounter, we extracted patient age and sex, documented diagnosis, number of medicines and antibiotics prescribed, injection prescribing, generic prescribing, and concordance with the UCG. No personally identifying information was collected.

### Outcome measures

The primary outcome was the proportion of prescriptions concordant with the UCG. A prescription was classified as concordant when the audit record indicated that treatment was consistent with UCG recommendations for the documented diagnosis.

Secondary outcomes were any antibiotic prescribed, defined as one or more antibiotics recorded for the encounter; number of medicines prescribed per encounter; a prescription containing only generic medicines, defined as an encounter in which all prescribed medicines were recorded using generic names; and any injection prescribed. Generic prescribing was therefore a prescription-level, rather than medicine-level, indicator. Prescribing from the essential medicines list was not included as an analytic outcome because comparable baseline and endline data could not be reproducibly reconstructed.

Exploratory analyses examined the distribution of medicines per encounter, antibiotic prescribing across diagnostic groups, and the proportion receiving two or more antibiotics among encounters in which at least one antibiotic was prescribed.

### Diagnostic classification and case mix

Diagnoses recorded at baseline and endline were harmonized into clinically coherent groups: respiratory infection, urinary tract infection, reproductive tract infection/sexually transmitted infection, gastrointestinal infection, febrile/infectious illness, acid-peptic disease, chronic medical conditions, skin/allergic disease, musculoskeletal/trauma, multiple diagnoses, and other conditions. Age, sex, and diagnostic group were used to characterize differences in case mix between the two assessment periods and were included as covariates in adjusted analyses.

### Data quality and management

Data were assessed for completeness, implausible values, identifier inconsistencies, discordance between antibiotic indicators and recorded antibiotic counts, and systematic repeated record sequences. Systematic repeated sequences identified in the endline dataset were excluded from the primary analysis, whereas isolated clinically identical prescriptions were retained because separate encounters could legitimately contain identical prescribing information. Where the recorded binary antibiotic indicator was inconsistent with the antibiotic count, the primary antibiotic outcome was defined consistently across both assessment periods using the recorded antibiotic count, with any value greater than zero classified as antibiotic prescribing.

### Statistical analysis

Age was summarized using mean (standard deviation [SD]) and median (interquartile range [IQR]); number of medicines per encounter using mean (SD); and categorical variables using frequencies and percentages. Baseline–endline differences in age, sex, and diagnostic case mix were described using standardized mean differences (SMDs) and descriptive *p* values. Mean age was compared using Welch’s *t* test; categorical variables using Pearson’s chi-square or Fisher’s exact test; and the overall diagnostic distribution using a Monte Carlo Fisher exact test.

For binary prescribing outcomes, we estimated baseline and endline proportions, crude absolute percentage-point differences, and 95% confidence intervals. Adjusted absolute differences were obtained from logistic regression using marginal standardization. Adjusted prevalence ratios (aPRs) and 95% confidence intervals were estimated using modified Poisson regression with robust standard errors. Models for UCG concordance, antibiotic prescribing, and generic prescribing included study period, continuous age, sex, and harmonized diagnostic group; sex and diagnostic group were modeled as categorical variables.

Number of medicines per encounter was analyzed using linear regression with heteroscedasticity-robust standard errors, adjusting for the same covariates, and reported as an adjusted mean difference with 95% confidence interval. Injection prescribing was compared using Fisher’s exact test without multivariable adjustment because events were sparse. Diagnosis-specific antibiotic prescribing and use of two or more antibiotics among antibiotic-treated encounters were exploratory and unadjusted.

All tests were two-sided; interpretation emphasized effect estimates and 95% confidence intervals. Analyses were conducted in R version 4.6.1 (R Foundation for Statistical Computing, Vienna, Austria).

### Ethical considerations

Administrative clearance for the quality improvement activity was obtained from the Ntungamo District Health Office and Itojo Hospital administration. The project underwent a public-health project-determination process by the Office of the Director of Science, U.S. Centers for Disease Control and Prevention (CDC) and was determined to be non-research, with the primary intent of improving public-health practice. The evaluation used retrospectively reviewed routine outpatient prescription and dispensing records; no personally identifying information was collected or included in the analytic dataset, and individual informed consent was not required for the record review. Data were handled confidentially and stored on password-protected computers accessible only to authorized project personnel.

## Results

### Prescription encounters and case mix

The baseline assessment included 350 systematically sampled outpatient prescription encounters. The raw endline dataset contained 198 records. Data-quality review identified 97 records within systematic repeated sequences, which were excluded from the primary analysis; isolated clinically identical prescriptions were retained. The final endline analytic sample therefore comprised 101 encounters.

Patients represented in the baseline and endline prescription samples were similar in age: mean age was 33.1 years (SD 22.2) at baseline and 31.2 years (SD 16.4) at endline (p=0.355), and 29.7% and 24.8%, respectively, were aged <18 years. The proportion of encounters involving female patients was higher at endline than at baseline (76.2% vs 62.9%; SMD 0.29; p=0.012). Diagnostic case mix also differed between periods (p<0.001), most notably for reproductive tract infection/sexually transmitted infection, multiple diagnoses, febrile/infectious illness, skin/allergic disease, and respiratory infection (Table 1).

**Table 1.**
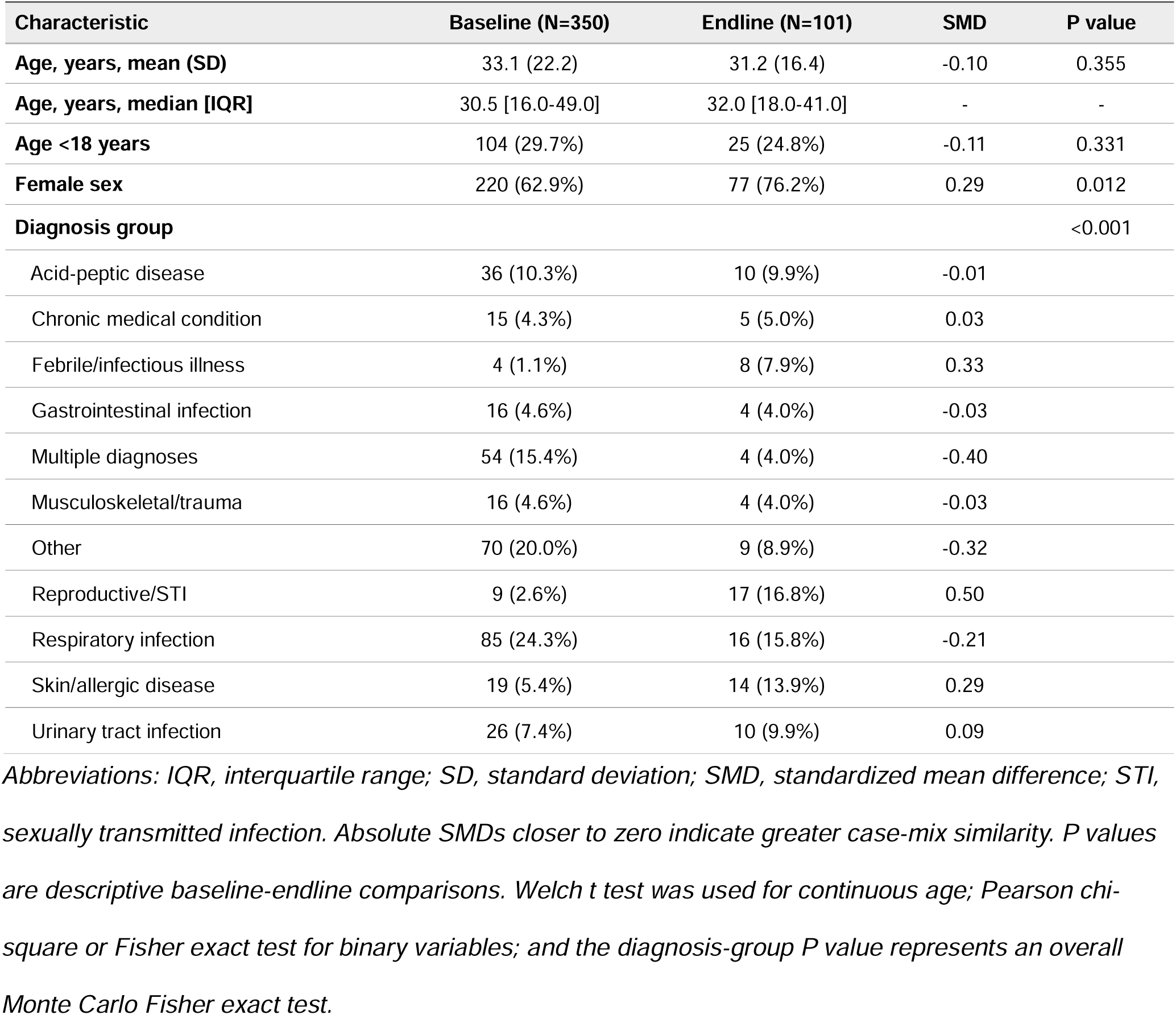
Characteristics and diagnostic case mix of outpatient prescription encounters.

### Root-cause findings and implementation of the QI change package

Root-cause analysis identified several modifiable contributors to inappropriate prescribing: limited access to UCG materials and essential medicines lists; inconsistent use of treatment guidelines; prescribing by staff outside the recognized clinician group; absence of routine prescription audit and feedback; and diagnostic limitations that encouraged empirical treatment, particularly antibiotic use. These findings informed a multifaceted change package implemented between May and November 2021, comprising prescriber education, provision of soft-copy PDF versions of the UCG to OPD clinicians, provision and quarterly updating of essential medicines lists, restriction of prescribing to recognized clinicians with signature verification at the dispensing unit, and periodic prescription audit and feedback (Table 2).

**Table 2.**
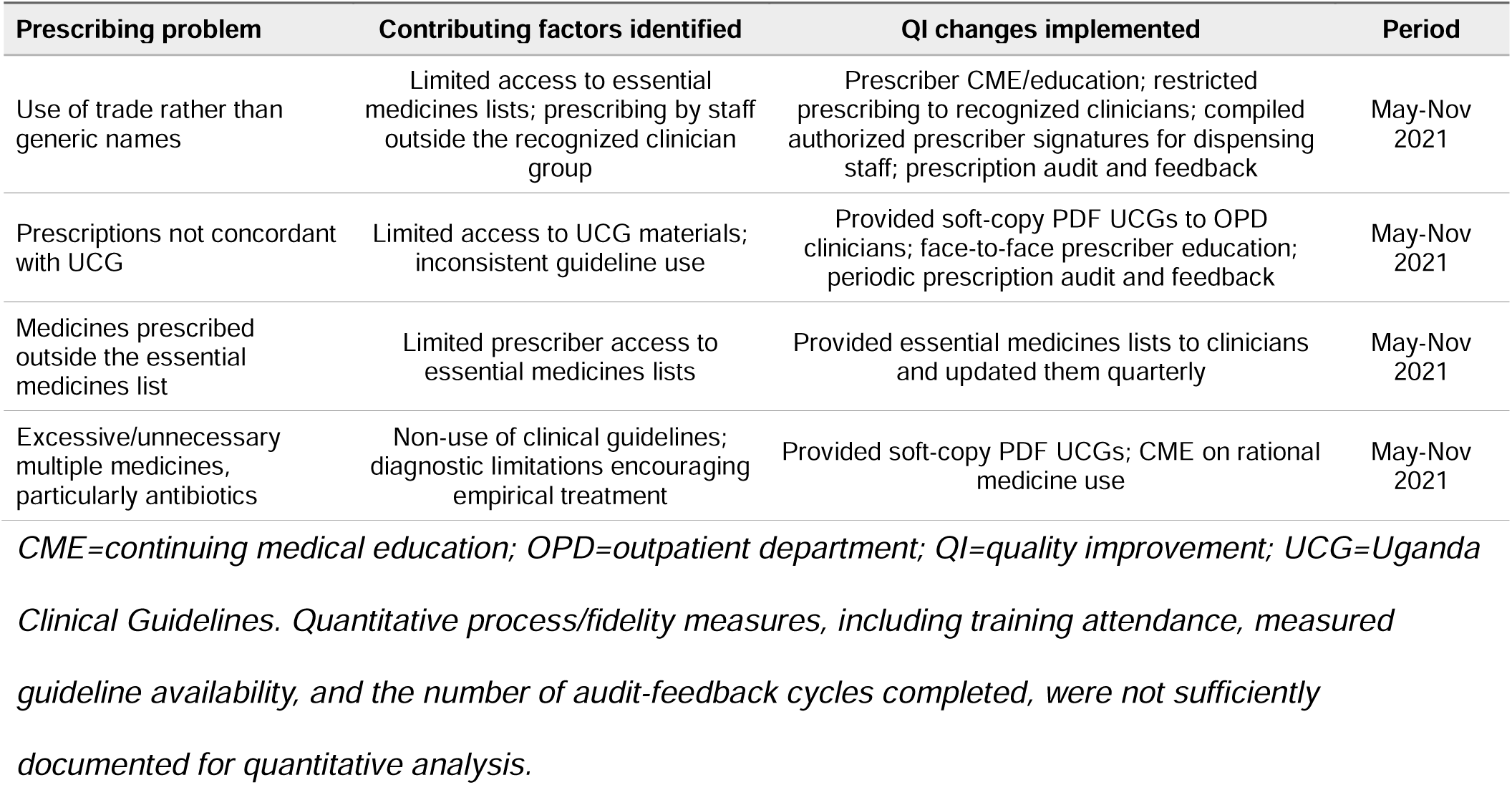
Prescribing problems, contributing factors, and QI changes implemented at Itojo Hospital.

### Guideline-concordant prescribing

The proportion of prescriptions concordant with the UCG, the primary outcome, was 43.7% (153/350) at baseline and 76.2% (77/101) at endline, corresponding to a crude absolute increase of 32.5 percentage points (95% CI 22.7 to 42.3). After adjustment for age, sex, and diagnostic group, UCG-concordant prescribing was 32.8 percentage points higher at endline (95% CI 21.9 to 43.8), with an adjusted prevalence ratio (aPR) of 1.82 (95% CI 1.51 to 2.20; p<0.001) (Fig 1). The magnitude of the baseline-to-endline difference was essentially unchanged after accounting for measured case-mix differences.

**Fig 1.**
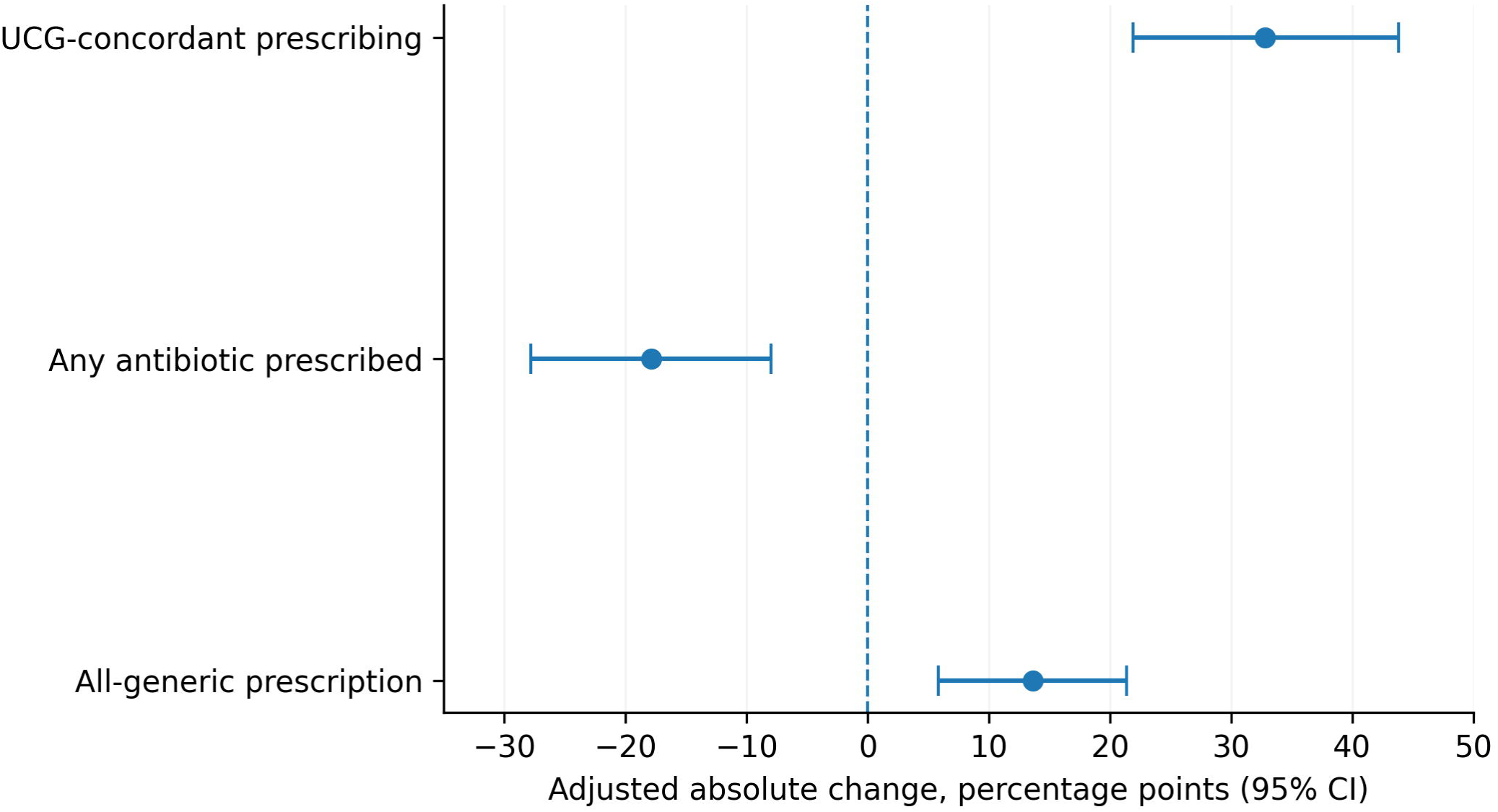
Adjusted baseline-to-endline changes in key prescribing indicators at Itojo Hospital, Uganda, 2021. Points represent case-mix-adjusted absolute percentage-point differences between endline and baseline; horizontal bars indicate 95% confidence intervals. Estimates were adjusted for age, sex, and harmonized diagnostic group. Positive values indicate a higher prevalence at endline and negative values indicate a lower prevalence at endline. UCG=Uganda Clinical Guidelines.

### Antibiotic prescribing

Any antibiotic was prescribed in 83.4% (292/350) of baseline encounters compared with 65.3% (66/101) at endline, a crude decrease of 18.1 percentage points (95% CI -28.1 to -8.0). The adjusted difference was -17.9 percentage points (95% CI -27.8 to -8.0), corresponding to an aPR of 0.80 (95% CI 0.70 to 0.91; p<0.001). Despite the lower prevalence at endline, antibiotics remained prescribed in almost two-thirds of outpatient encounters.

Among encounters in which at least one antibiotic was prescribed, the proportion containing two or more antibiotics did not materially differ, at 43.5% (127/292) at baseline and 48.5% (32/66) at endline (crude change, +5.0 percentage points; 95% CI -8.3 to 18.3; p=0.461).

### Number of medicines prescribed

The mean number of medicines per encounter was 3.35 (SD 1.02) at baseline and 2.66 (SD 0.94) at endline, a crude mean difference of -0.68 medicines per encounter (95% CI -0.90 to -0.47; p<0.001). After adjustment for age, sex, and diagnostic group, the mean difference was -0.61 medicines per encounter (95% CI -0.79 to -0.43).

The distribution of medicine counts shifted toward fewer medicines at endline (Fig 2). At baseline, 12% of encounters contained five or more medicines and 27% contained four medicines, compared with approximately 4% and 9%, respectively, at endline. Prescriptions containing one or two medicines accounted for approximately 18% of baseline encounters and 39% of endline encounters. Three medicines remained the most common prescription size in both periods (43% vs 49%).

**Fig 2.**
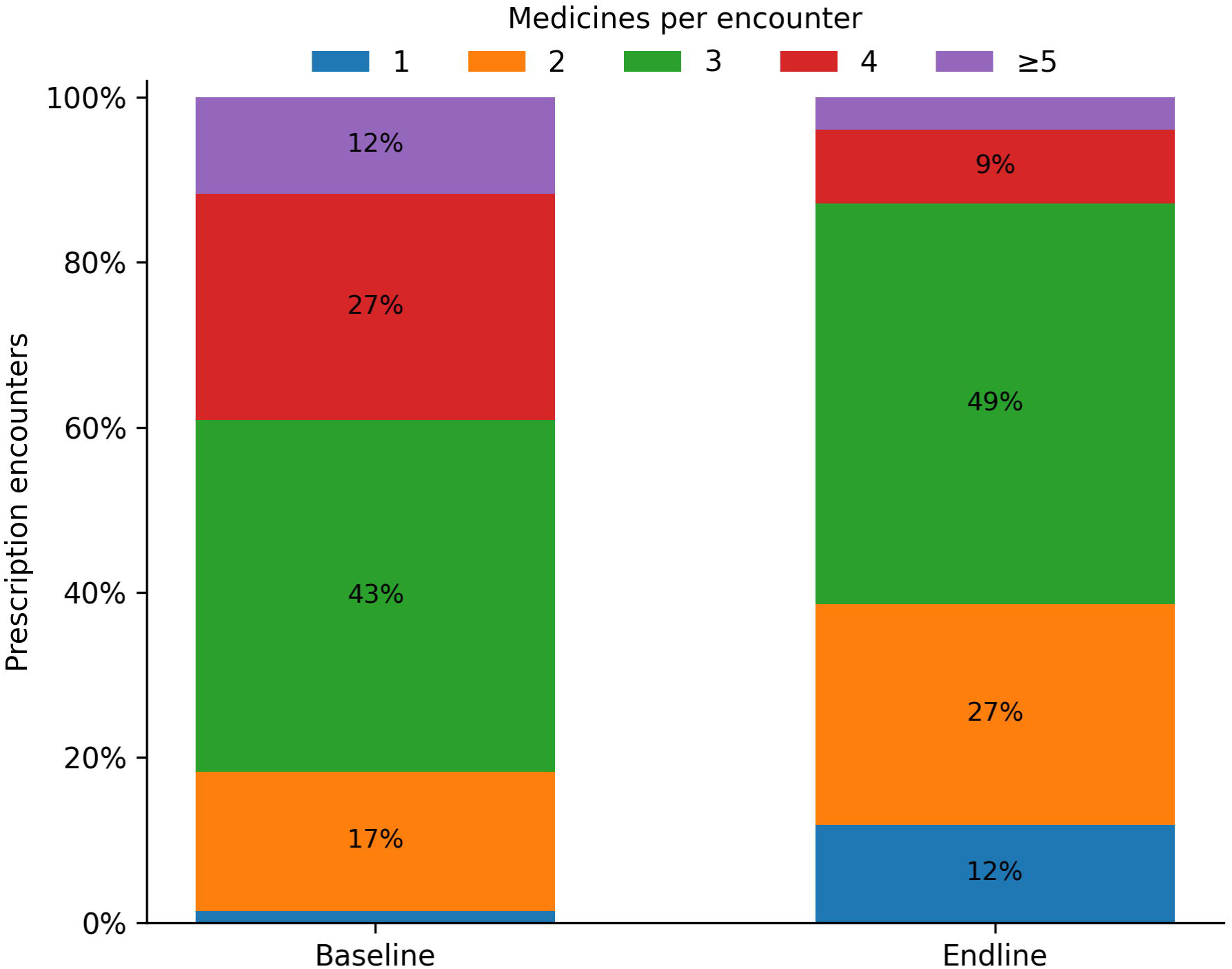
Medicines prescribed per outpatient encounter at baseline and endline, Itojo Hospital, Uganda. Bars show the percentage of prescription encounters containing one, two, three, four, or five or more medicines. Mean medicines per encounter were 3.35 at baseline and 2.66 at endline.

### Generic and injection prescribing

Prescriptions containing only medicines recorded by generic name increased from 78.6% (275/350) at baseline to 94.1% (95/101) at endline. The crude increase was 15.5 percentage points (95% CI 9.2 to 21.8), and the adjusted increase was 13.6 percentage points (95% CI 5.8 to 21.4; aPR 1.16, 95% CI 1.06 to 1.26; p<0.001)

Injection prescribing was uncommon at both assessments and showed little evidence of change: 3.1% (11/350) of baseline encounters and 4.0% (4/101) of endline encounters included an injection (absolute change +0.8 percentage points; 95% CI -3.4 to 5.0; p=0.753). Given the small number of injection encounters, no adjusted model was fitted. Table 3 summarizes the primary and secondary prescribing outcomes.

**Table 3.** Baseline-to-endline changes in rational prescribing indicators.

| Outcome | Baseline | Endline | Crude change (95% CI) | Adjusted change (95% CI) | Adjusted relative effect (95% CI) | P value |
| --- | --- | --- | --- | --- | --- | --- |
| Prescription concordant with UCG | 153/350 (43.7%) | 77/101 (76.2%) | +32.5 pp (22.7 to 42.3) | +32.8 pp (21.9 to 43.8) | 1.82 (1.51 to 2.20) | <0.001 |
| Any antibiotic prescribed | 292/350 (83.4%) | 66/101 (65.3%) | -18.1 pp (-28.1 to -8.0) | -17.9 pp (-27.8 to -8.0) | 0.80 (0.70 to 0.91) | <0.001 |
| Number of medicines per encounter, mean (SD) | 3.35 (1.02) | 2.66 (0.94) | -0.68 (-0.90 to -0.47) | -0.61 (-0.79 to -0.43) | - | <0.001 |
| Prescription containing only generic medicines | 275/350 (78.6%) | 95/101 (94.1%) | +15.5 pp (9.2 to 21.8) | +13.6 pp (5.8 to 21.4) | 1.16 (1.06 to 1.26) | <0.001 |
| Injection prescribed | 11/350 (3.1%) | 4/101 (4.0%) | +0.8 pp (-3.4 to 5.0) | Not adjusted | - | 0.753 |

*Adjusted binary estimates are marginal standardized percentage-point differences from logistic models; adjusted relative effects are prevalence ratios from modified Poisson regression with robust standard errors. Models adjust for age, sex, and harmonized diagnostic group. Number of medicines was analyzed using robust linear regression. Injection use was sparse and was not adjusted. Generic prescribing is a prescription-level measure in which all medicines on the prescription were recorded using generic names. aPR=adjusted prevalence ratio; pp=percentage points; UCG=Uganda Clinical Guidelines.*

### Antibiotic prescribing by diagnosis

Antibiotic prescribing varied substantially across diagnostic groups at both assessments (S1 Fig). The direction and magnitude of baseline-to-endline differences were not uniform across diagnoses: lower endline prescribing was apparent in several groups, including skin/allergic disease, acid-peptic disease, chronic medical conditions, and the heterogeneous other category, whereas prescribing remained high for urinary tract and respiratory infections. Some diagnostic groups contained few endline encounters.

## Discussion

This facility-led QI initiative was accompanied by substantial improvements in several measures of prescribing quality. Guideline-concordant prescribing increased markedly, antibiotic use declined, medicine burden per encounter was reduced, and generic prescribing became more consistent. The magnitude of change in the primary outcome remained substantial after adjustment for differences in age, sex, and diagnostic case mix. Nevertheless, antibiotic prescribing remained common at endline and the proportion of antibiotic-treated encounters receiving two or more antibiotics was unchanged. The findings therefore suggest meaningful progress in rational prescribing, but incomplete improvement in antimicrobial use.

The marked increase in guideline-concordant prescribing addresses a problem that extends beyond a single facility. Across Uganda, adherence to national treatment guidance has remained suboptimal, particularly for antibiotic prescribing. Although our outcome assessed concordance of the overall prescription rather than antibiotic appropriateness alone, the pattern is consistent with previous evidence: UCG concordance was only 34.2% among nearly 10,000 antibiotic prescriptions in lower-level facilities in southwestern Uganda [5] and approximately 30% in a national survey of 13 hospitals[11]. This broader context highlights the importance of implementation strategies that move clinical guidelines from policy documents into routine prescribing workflows.

The observed improvement is plausibly related to the multifaceted nature of the change package, although the contribution of individual components cannot be determined. In addition to prescriber education, clinicians received soft-copy PDF UCGs, prescribing roles were clarified, signature verification strengthened accountability, and audit and feedback were introduced. These changes directly addressed barriers identified during the root-cause analysis. Evidence from low- and lower-middle-income settings supports this approach: most interventions to improve antibiotic-guideline adherence are multifaceted and commonly combine education with monitoring, review, or feedback [12], while provider education appears more effective when paired with enabling and supportive strategies[13]. This suggests that sustained improvement in prescribing is more likely when educational and system-level barriers are addressed together.

Our findings are broadly consistent with Uganda’s SPARS experience, which demonstrates the value of linking measurement with supervision, feedback, and capacity building rather than relying on data collection alone. Across more than 1,200 facilities, SPARS was associated with improvements in several appropriate-medicine-use indicators, although performance gains varied across indicators and levels of care[4]. The smaller, locally led Itojo initiative similarly suggests that routine prescribing information can support meaningful improvement when it is translated into targeted feedback and practical changes in clinical workflow.

The decline in antibiotic prescribing is encouraging, but the 65.3% endline prevalence underscores that antibiotic use remained a major quality-of-care concern. Similar patterns have been documented elsewhere in Uganda, with antibiotic prescribing reported in 66.2% of outpatient encounters at a secondary-care hospital in western Uganda [6], and in roughly 80% of respiratory tract infection encounters in eastern Uganda [7]. Across African primary-care settings, the median prevalence was 46.8% [3], although marked heterogeneity across populations and facilities limits direct comparison. The Itojo findings, therefore, suggest improvement in overall antibiotic use, but not attainment of a satisfactory stewardship endpoint.

Continued high antibiotic use likely reflects determinants beyond prescriber knowledge. The QI team identified diagnostic limitations and empirical treatment as important contributors, consistent with qualitative evidence from Uganda showing that antibiotic decisions are shaped by diagnostic uncertainty, medicine availability, clinician judgement, patient expectations, and broader norms of care [14]. These persistent influences may explain why overall antibiotic prescribing declined while the proportion of antibiotic-treated encounters receiving multiple antibiotics did not. Future improvement efforts may therefore need to pair prescribing feedback with diagnostic stewardship and more targeted review of antibiotic indication, selection, and combination therapy.

The residual antibiotic burden has implications beyond the facility. Antimicrobial resistance remains a major cause of morbidity and mortality in Africa, with approximately 1.05 million deaths associated with bacterial AMR and 250,000 attributable deaths estimated in the WHO African Region in 2019[15]. Uganda’s national AMR action plan prioritizes antimicrobial stewardship, appropriate prescribing, improved diagnostics, training, and surveillance[16]. Facility-level QI may therefore provide a practical mechanism for translating these national priorities into routine clinical practice, particularly in district hospitals where dedicated antimicrobial-stewardship expertise may be limited.

The reduction in medicine burden suggests a broader shift in prescribing practice. Mean medicines per encounter decreased from 3.35 to 2.66, and high-count prescriptions became less frequent. The baseline mean was comparable to estimates reported in the WHO African Region and at another secondary-care hospital in western Uganda[3,6]. Because medicine requirements depend on clinical need, fewer medicines should not be equated automatically with better care. In this context, however, the simultaneous improvement in UCG concordance makes it more plausible that prescribing became more selective and guideline-aligned.

Generic-name prescribing also improved, with a greater proportion of encounters containing only medicines recorded by generic name. Because this definition applies to the entire prescription, it should not be directly benchmarked against the conventional WHO indicator based on individual medicines. Even so, the within-facility change suggests more consistent generic prescribing. Injection use remained low and essentially unchanged at 3–4%, likely because baseline performance was already favorable and offered little room for further improvement. Similar floor effects have been observed in Uganda’s SPARS experience[4].

From a health-system perspective, the intervention was designed to fit within existing facility structures rather than operate as a parallel program. Electronic access to the UCG and use of routine prescribing data for audit and feedback may improve practicality in resource-constrained settings. However, the present evaluation did not assess implementation costs or cost-effectiveness, including staff time, training, supervision, and digital access. Because audit-and-feedback interventions vary in both effect and resource requirements across settings[17], decisions about wider implementation should be informed by economic evaluation as well as evidence on medicine expenditure, stock-outs, and patient outcomes.

### Limitations

Several limitations should inform interpretation. First, the study design limits causal inference. This was a single-facility, uncontrolled before-and-after evaluation, so secular trends or concurrent changes in staffing, medicine availability, or disease patterns may have influenced the findings. Although analyses adjusted for age, sex, and diagnostic group, residual confounding remains possible. Second, sampling and routine data limitations may have affected comparability and precision. The baseline covered three months, whereas endline covered three weeks, and repeated endline sequences were excluded during data cleaning, leaving 101 analyzable encounters. Outcome classification also depended on routine documentation, which may have been incomplete. Third, implementation fidelity was not quantified. We lacked complete data on training coverage, actual guideline use, and audit-feedback cycles, and therefore could not assess intervention dose or the contribution of individual components. Patient outcomes, medicine stock-outs, antimicrobial resistance, adverse events, and costs were also not measured.

Despite these limitations, the study used systematic sampling, evaluated multiple prescribing outcomes, adjusted for measured case-mix differences, and reported both absolute and relative estimates. The QI package was also developed from locally identified root causes and implemented through existing hospital structures, supporting its practical relevance.

## Conclusions

This facility-led quality improvement initiative was associated with substantial improvements in several measures of rational prescribing, including greater concordance with the Uganda Clinical Guidelines, lower antibiotic use, fewer medicines per encounter, and more consistent generic prescribing. However, antibiotic prescribing remained common, suggesting that important drivers of antimicrobial use persisted despite the intervention package. Sustained improvement will likely require continued audit and feedback, stronger diagnostic stewardship, and repeated quality-improvement cycles. Further evaluation should assess whether these gains are maintained over time and can be adapted across similar district-hospital settings.

## Supporting information

S1 Fig. Antibiotic prescribing by diagnostic group at baseline and endline

## Data Availability

All data underlying the findings reported in the manuscript are provided in S1 Data. The data dictionary is provided as S2 Data, and full adjusted regression outputs as S1 Table.

## Acknowledgments

We thank the Itojo Hospital quality improvement team, hospital administration, clinicians, pharmacy and dispensing staff, laboratory staff, and records personnel for their participation in identifying prescribing gaps and implementing the improvement activities. We also acknowledge the Ntungamo District Health Office for administrative support and the Uganda Public Health Fellowship Program and Uganda National Institute of Public Health for technical and administrative support during the project.

## Author contributions (CRediT)

RM: Conceptualization; Data curation; Formal analysis; Investigation; Methodology; Project administration; Supervision; Visualization; Writing – original draft; Writing – review & editing.

SN: Data curation; Formal analysis; Writing – original draft; Writing – review & editing.

BK: Methodology; Supervision; Writing – review & editing.

LB: Methodology; Writing – review & editing.

ARA: Conceptualization; Supervision; Writing – review & editing; Funding acquisition

## Data Availability Statement

The datasets used and analyzed during this study are available from the corresponding author upon reasonable request.

## Supporting information captions

S1 Table. Full adjusted regression model output. Coefficients, robust standard errors, 95% confidence intervals, p values, and exponentiated estimates where applicable are provided for the logistic, modified Poisson, and robust linear regression models used in the main analysis.

S1 Fig. Antibiotic prescribing by diagnostic group at baseline and endline, Itojo Hospital, Uganda, 2021. Points show the crude proportion of encounters within each diagnostic group in which at least one antibiotic was prescribed. Lines connect baseline and endline estimates. The analysis was exploratory; diagnostic groups with fewer than four encounters in either period are omitted from the figure.

