## Supplementary figures and images for "Improving rational medicine prescribing at a public district hospital in southwestern Uganda, January–November 2021: a quality improvement initiative"

### S1 Fig. Antibiotic prescribing by diagnostic group at baseline and endline

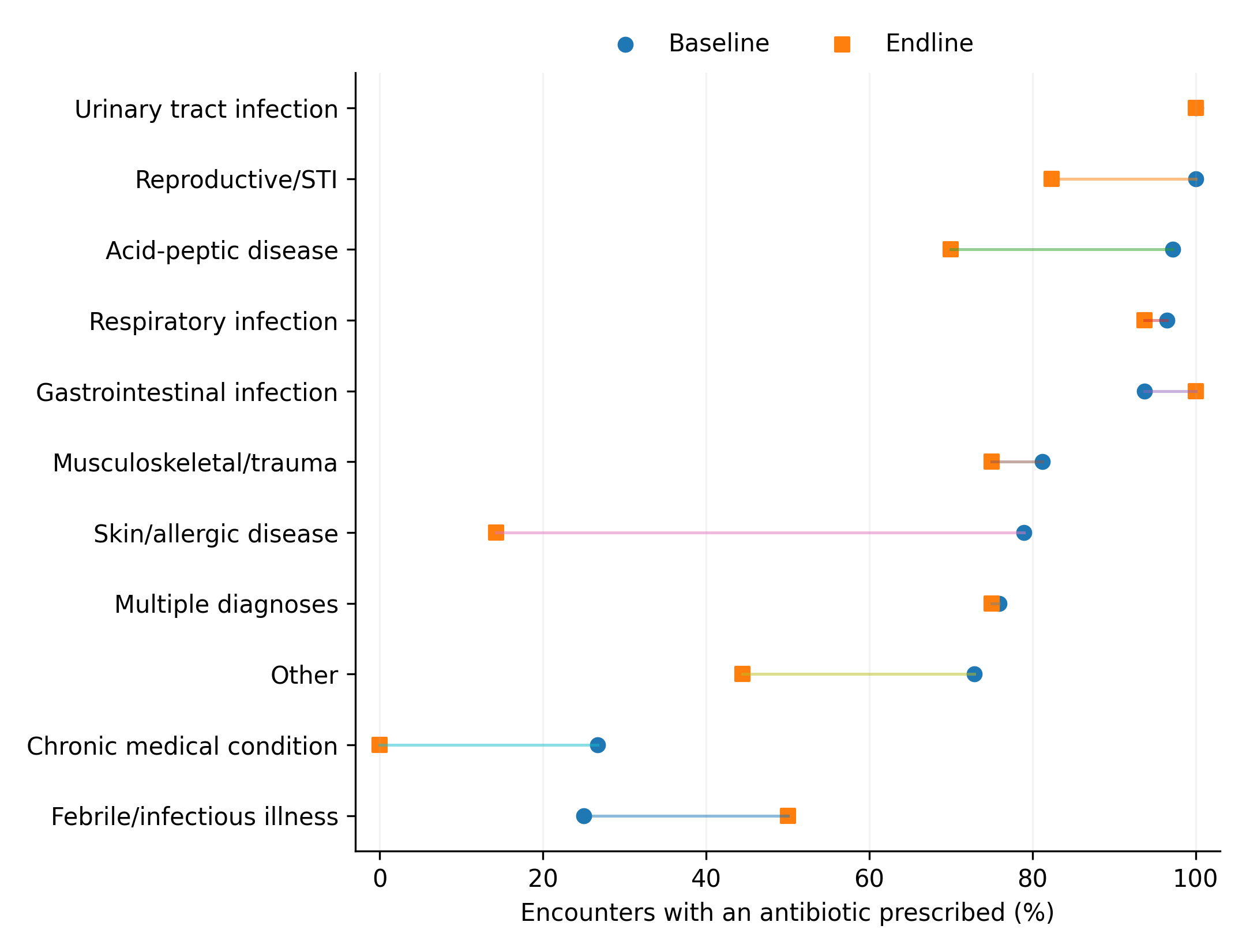
